# Weak anti-SARS-CoV-2 antibody response after the first injection of an mRNA COVID-19 vaccine in kidney transplant recipients

**DOI:** 10.1101/2021.03.08.21252741

**Authors:** Ilies Benotmane, Gabriela Gautier-Vargas, Noelle Cognard, Jerome Olagne, Francoise Heibel, Laura Braun-Parvez, Jonas Martzloff, Peggy Perrin, Bruno Moulin, Samira Fafi-Kremer, Sophie Caillard

**Affiliations:** Department of Nephrology and Transplantation, University Hospital, Strasbourg, France; Fédération de Médecine Translationnelle (FMTS), Strasbourg, France; Department of virology, University Hospital, Strasbourg, France

**Keywords:** kidney transplant recipient, COVID-19 vaccine

## Abstract

Data concerning the anti-SARS-CoV-2 antibody response after mRNA COVID-19 vaccine in kidney transplant recipients (KTRs) are currently lacking. Here, we sought to examine this issue by analyzing the serological response observed in 241 KTRs after a first vaccine injection. Our results indicate that KTRs have a weak anti-SARS-CoV-2 antibody response, ultimately resulting in a low seroconversion rate (26/241, 10.8%). This phenomenon likely stems from a high immunosuppression burden in this clinical population.

---

International recommendations on COVID-19 vaccine distribution have given priority to immunocompromised patients – including kidney transplant recipients (KTRs).^1,2^ Unfortunately, this guidance has been released without inclusion of this clinical population in vaccine clinical trials. In an effort to shed light on the efficacy and safety of an mRNA COVID-19 vaccine in KTRs, this preliminary study was undertaken to investigate the anti-SARS-CoV-2 antibody response after the first injection. The protocol was reviewed and approved by the local Institutional Review Board (approval number: DC-2013–1990).

We examined 242 KTR who received the first injection of the Moderna mRNA-1273 vaccine (100 µg) at the Strasbourg University Hospital (France) between January 21 and 28, 2021. All had a negative history for COVID-19 and tested negative for anti-SARS-CoV-2 antibodies on the day of the first injection. The anti-SARS-CoV-2 antibody response was assessed at 28 days post-injection using the ARCHITECT IgG II Quant test (Abbott, Abbott Park, IL, USA), with titers >50 AU/mL being considered as positive.

One patient developed mild symptomatic COVID-19 seven days after injection and only 26 (10.8%) KTRs had a positive serology at 28 days post-injection. The median IgG titer was 224 AU/mL (interquartile range: 76-496 AU/mL). Patients who seroconverted had longer time from transplantation, received less immunosuppression, and had a better kidney function (Table 1).

**Table 1.** Characteristics of kidney transplant recipients according to serological response after the first dose of the Moderna mRNA-1273 vaccine

|  | Entire cohort<br>(n=241*) | SARS-CoV-2<br>seronegative<br>patients<br>(n=215) | SARS-CoV 2<br>seropositive<br>patients (n=26) | p | Missing<br>data |
| --- | --- | --- | --- | --- | --- |
| **Age (years)** | 57.7 (49.3-67.6) | 57.7 (49.6-67.7) | 58.4 (43.3--66.9) | 0.51 | 0 |
| **Male sex** | 156 (64.7%) | 142 (66.1%) | 14 (53.9%) | 0.28 | 0 |
| **BMI (kg/m<sup>2</sup>)** | 25.7 (22.6-29.5) | 25.7 (22.8-29.4) | 26.4 (21.9-29.9) | 0.73 | 2 |
| **Time from kidney transplantation (years)** | 6.4 (2.9-13) | 5.8 (2.8-11.9) | 15.4 (8.6-25.9) | <0.001 | 2 |
| **First transplantation** | 202 (83.8%) | 176 (81.8%) | 26 (100%) | 0.01 | 0 |
| **Deceased donor** | 192 (79.6%) | 172 (80%) | 20 (76.9%) | 0.8 | 0 |
| **ABO Group** |  |  |  | 0.02 | 2 |
| **O** | 94 (39.3%) | 82 (38.5%) | 12 (46.2%) |  |  |
| **A** | 101 (42.3%) | 93 (43.7%) | 8 (30.8%) |  |  |
| **B** | 30 (12.6%) | 29 (13.6%) | 1 (3.9%) |  |  |
| **AB** | 14 (5.9%) | 9 (4.2%) | 5 (19.2%) |  |  |
| **Induction treatment** |  |  |  | 0.001 | 7 |
| **Anti-thymocyte globulin** | 138 (59.5%) | 127 (60.8%) | 11 (47.8%) |  | 9 |
| **Anti-CD25** | 88 (37.9%) | 80 (38.3%) | 8 (34.8%) |  |  |
| **No induction** | 6 (2.6%) | 2 (1%) | 4 (17.4%) |  |  |
| **CNI** |  |  |  | 0.06 | 0 |
| **Tacrolimus** | 133 (55.2%) | 124 (57.7%) | 9 (34.6%) |  |  |
| **Ciclosporin** | 82 (34%) | 69 (32.1%) | 14(50%) |  |  |
| **No CNI** | 26 (10.8%) | 22 (10.2%) | 4 (15.4%) |  |  |
| **MMF/MPA** | 191 (79.3%) | 177 (82.3%) | 14 (53.9%) | 0.002 | 0 |
| **Imurel** | 7 (2.9%) | 4 (1.86%) | 3 (11.5%) | 0.03 | 0 |
| **mTOR inhibitors** | 35 (14.5%) | 32 (14.9%) | 3 (11.6%) | 1 | 0 |
| **Steroids** | 142 (58.9%) | 133 (61.9%) | 9 (34.6%) | 0.01 | 0 |
| **Belatacept** | 9 (3.8%) | 9 (4.2%) | 0 | 0.26 | 0 |
| **eGFR (mL/min/1.73m<sup>2</sup>)** | 51.6 (38.1-68) | 51 (37.9-66.5) | 64.9 (39.9-72.2) | 0.08 | 0 |
| **Serum creatinine (μmol/L)** | 118 (99-158) | 120 (101-159) | 104 (85-134) | 0.05 | 0 |

\*The patient who developed COVID-19 after the first injection was excluded from the analysis.
Continuous variables are presented as medians (interquartile ranges), whereas categorical variables are presented as counts (percentages).
Abbreviations: BMI, body mass index; CNI, calcineurin inhibitors; eGFR, estimated glomerular filtration rate; MMF, mycophenolate mofetil; MPA, mycophenolic acid

In summary, the burden of immunosuppression may induce a weak anti-SARS-CoV-2 antibody response in KTRs after the first injection of an mRNA COVID-19 vaccine. This finding is strikingly different compared with immunocompetent subjects – who invariably seroconverted after the first injection.^3,4^ We highlight the need not to delay the second vaccine injection in immunocompromised patients. Close surveillance is also recommended to discuss the opportunity of a third dose in less responsive patients.

## Data Availability

Data supporting the findings from this study are available from the corresponding author upon reasonable request.

